# Neurogenic dysphagia as an independent driver of hospital length of stay and costs: *a Bayesian analysis with geriatric stratification and intervention simulation*

**DOI:** 10.64898/2026.04.08.26350417

**Authors:** Cornelius J. Werner, Tareq Meyer, João Pinho, Bettina Mall, Jörg B. Schulz, Beate Schumann-Werner

## Abstract

**Purpose:** Neurogenic dysphagia is prevalent in neurological inpatients and associated with adverse outcomes, yet its independent economic impact after adjustment for frailty and functional status remains poorly quantified. We aimed to estimate the independent effect of dysphagia on hospital length of stay (LOS) and costs, to test whether this effect differs between geriatric and non-geriatric patients, and to quantify the probability and magnitude of cost savings from improvements in swallowing function.

**Methods:** We analysed 10,375 neurological inpatient cases (2021–2024) at a German university hospital. Dysphagia was defined by fiberoptic endoscopic evaluation of swallowing (FEES) or ICD-10 R13 coding (n = 1,382; 13.3%). Bayesian Gamma-log regression with informative priors from historical data and published literature was used to model LOS and total case costs (German DRG), adjusted for age, sex, Hospital Frailty Risk Score (HFRS, R13-adjusted), self-care index („Selbstpflege-Index“, SPI), stroke status, and emergency admission. A geriatric cohort was defined as age ≥70 and adjusted HFRS ≥5 (n = 2,053; 19.8%). Posterior predictive simulation estimated cost savings for hypothetical improvements of 1–3 points on the Functional Oral Intake Scale (FOIS).

**Results:** After comprehensive adjustment, dysphagia was independently associated with 46.5% longer LOS (posterior ratio 1.465; 95% credible interval [CrI] 1.397–1.537) and 28.2% higher total case costs (ratio 1.282; CrI 1.213–1.354). The dysphagia × geriatric interaction was small but credible and ran in opposite directions: slightly attenuated for LOS (interaction ratio 0.908, CrI 0.837–0.986) but slightly amplified for costs (1.096, CrI 1.012–1.185), consistent with complexity-driven DRG grouping in geriatric patients. The absolute economic burden remained larger in the geriatric cohort due to higher baseline costs. In the geriatric cohort, a one-point FOIS improvement yielded a 74.3% posterior probability of LOS-based savings (mean €555/case); at three points, this rose to 84.2% (mean €1,115/case). The direct cost model confirmed high benefit probabilities from the payer’s perspective (82.6% at δFOIS = 3).

**Conclusions:** Neurogenic dysphagia is an independent and substantial driver of hospital LOS and costs in neurological inpatients, even after adjustment for frailty and functional status. The proportional effect on costs is slightly larger in geriatric patients, while the LOS effect is slightly smaller, consistent with the mechanics of the G-DRG system. Bayesian simulation indicates that improvements in swallowing function carry a high probability of generating cost savings, supporting the characterisation of dysphagia as a modifiable economic target with particular relevance to geriatric neurology.

## Introduction

Neurogenic dysphagia is among the most common and consequential complications of neurological disease. It affects 37–78% of acute stroke patients depending on the method of assessment [1], up to 82% of patients with Parkinson’s disease when tested objectively [2], and 27–57% of patients with dementia [3,4]. The clinical consequences extend far beyond impaired oral intake: a recent meta-analysis of 67 studies demonstrated that dysphagia confers a nearly tenfold increase in the risk of aspiration pneumonia (OR 9.60; 95% CI 5.75–16.04) in acute stroke [5], and is independently associated with malnutrition, dehydration, prolonged hospitalisation, and increased mortality [1,6,7].

The prevalence of oropharyngeal dysphagia increases markedly with age, affecting 30–40% of independently living older adults and reaching 68.9% in acute geriatric settings [8,9] In recognition of its high prevalence and systemic impact, the European Society for Swallowing Disorders and the European Union Geriatric Medicine Society jointly designated oropharyngeal dysphagia as a geriatric syndrome [8]. However, recent evidence indicates that the age–dysphagia association is fully mediated through functional decline rather than chronological aging per se (Werner CJ et al., submitted), underscoring the importance of distinguishing presbyphagia from pathological dysphagia [10].

Despite its clinical significance, the economic impact of neurogenic dysphagia remains incompletely understood. A meta-analysis of 23 studies showed that oropharyngeal dysphagia increases hospital length of stay by approximately 3 days (95% CI 2.7–3.3) and adds an estimated 40% to healthcare costs [11]. In the largest available study, Patel et al. [12] analysed nearly 88 million US inpatient records and reported a 76% increase in length of stay and $6,243 in additional costs attributable to dysphagia. In the German context, Labeit et al. [13] demonstrated that severe post-stroke dysphagia (FEDSS = 6) predicted €2,554 in additional health-insurance costs. However, the available evidence has three important limitations. First, most cost estimates are either unadjusted or adjusted only for basic demographics, without accounting for frailty or functional status, leaving open the question of whether dysphagia is an independent cost driver or merely a marker of overall disease severity. Second, to our knowledge, no study has explicitly tested whether the economic impact of dysphagia differs between geriatric and non-geriatric patients within the same population. Third, while clinical trials have demonstrated that dysphagia interventions can improve functional outcomes [14,15], economic evaluations of such interventions remain scarce [16], and no study has, to our knowledge, probabilistically quantified the potential cost savings associated with specific improvements in swallowing function.

Bayesian methods offer particular advantages for health economic analyses, including direct probabilistic statements about cost differences, principled incorporation of prior evidence, and full posterior distributions enabling intuitive clinical interpretation [17,18]. The Gamma distribution with log link has been established as the preferred model for right-skewed healthcare cost data [19–21], and can be implemented with informative priors using the brms R package [22,23].

The present study addresses these gaps by analysing the impact of neurogenic dysphagia on hospital length of stay and costs in a large cohort of neurological inpatients (N = 10,375) at a German university hospital, using Bayesian Gamma-log regression with informative priors. We pursued three objectives: (1) to estimate the independent effect of dysphagia on length of stay and costs after comprehensive adjustment for frailty (Hospital Frailty Risk Score), functional status (self-care index), and other confounders; (2) to test whether this effect differs between geriatric (age ≥70 and HFRS ≥5) and non-geriatric patients; and (3) to use Bayesian posterior predictive simulation to quantify the probability and magnitude of cost savings associated with improvements in functional swallowing status, as measured by the Functional Oral Intake Scale (FOIS) [24].

## Materials and Methods

### Study design and setting

This retrospective single-centre cohort study analysed routine clinical and administrative data from the Department of Neurology, University Hospital RWTH Aachen (Uniklinik RWTH Aachen), Germany. The study included all neurological inpatient cases discharged between January 1, 2021 and December 31, 2024. The study was approved by the institutional ethics committee of the Medical Faculty of RWTH Aachen University (EK088/17); the requirement for informed consent was waived given the retrospective, anonymised design. Reporting follows the Strengthening the Reporting of Observational Studies in Epidemiology (STROBE) guidelines for cohort studies.

### Participants

From an initial dataset of 15,390 inpatient cases, we applied sequential exclusion criteria (Figure 1). Cases with a length of stay (LOS) of zero days or missing LOS data were excluded (n = 467), as were cases with documented mechanical ventilation during the hospital stay (n = 789), defined as ventilation duration > 0 hours. Finally, cases with missing values for any mandatory covariate (age, sex, Hospital Frailty Risk Score [HFRS], or Self-Care Index [SPI]) were excluded (n = 3,759; predominantly due to missing SPI documentation). The final analytical sample comprised N = 10,375 cases.

**Figure 1.**
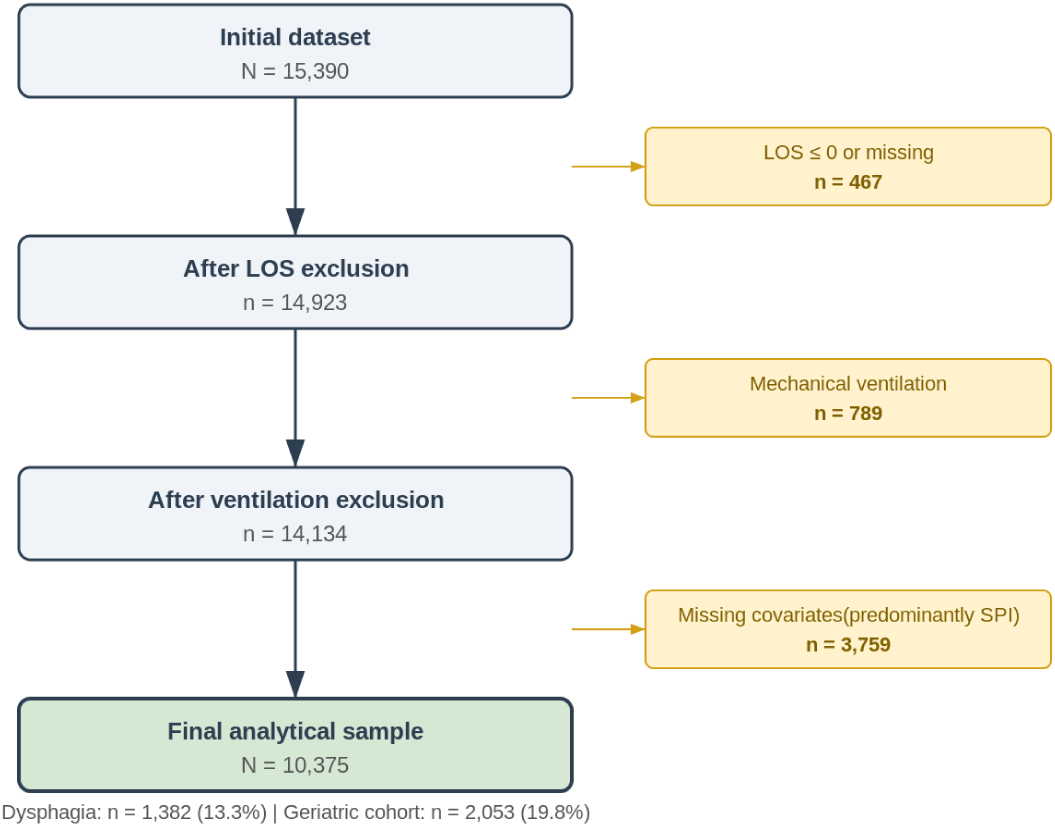
Study flow diagram showing patient selection and exclusion criteria.

### Dysphagia assessment and classification

Dysphagia status was determined using a composite criterion. Patients were classified as having documented dysphagia if they had either (a) undergone fiberoptic endoscopic evaluation of swallowing (FEES) during the index admission (FEES indicator = 1 in the clinical documentation system) resulting in diagnosis of dysphagia, and/or (b) received an ICD-10 diagnosis code of R13 (dysphagia) at discharge. This composite definition captures both instrumentally confirmed dysphagia and clinically coded dysphagia, reflecting real-world documentation practice. All remaining patients were classified as non-dysphagic. In total, 1,382 cases (13.3%) met the dysphagia criterion.

For patients who underwent FEES, dysphagia severity was quantified using the Functional Oral Intake Scale (FOIS), a 7-point ordinal scale ranging from 1 (nothing by mouth) to 7 (total oral diet with no restrictions) [24]. FOIS scores were available for 1,076 of 1,382 dysphagic patients (77.9%). Non-dysphagic patients were assigned a FOIS score of 7 (full oral intake), mirroring clinical practice. For the intervention simulation models (see below), only patients with documented FOIS scores were included. Missing FOIS values among dysphagic patients were handled by complete-case exclusion from the FOIS-based models.

### Outcome variables

Two co-primary outcomes were analysed: (1) length of stay (LOS), defined as the number of calendar days from admission to discharge, and (2) total case costs, defined as the actual Diagnosis-Related Group (DRG) revenue billed for each case (in Euros). Both outcomes represent positive, right-skewed, continuous variables. No cost imputation was performed; all cost figures reflect realised institutional DRG revenue.

### Covariates

The following covariates were included in all adjusted models: age (continuous, standardised), sex (male/female), stroke status (binary; derived from ICD-10 diagnostic group coding), emergency admission (binary), Hospital Frailty Risk Score (HFRS; continuous, standardised), and Self-Care Index at admission (SPI; continuous, standardised).

The Hospital Frailty Risk Score (HFRS) was calculated from ICD-10 diagnostic codes according to Gilbert et al. [25]. To avoid circularity between the dysphagia predictor and the frailty covariate, the HFRS was adjusted by subtracting the R13 (dysphagia) component weight of 0.8 for all patients carrying an R13 diagnosis code, consistent with the approach described in a separate analysis of dysphagia and functional status from the same institution (Werner CJ et al., submitted). This adjustment affected 1,284 patients in the final sample.

The Self-Care Index (SPI) is derived from the ePA-AC (ergebnisorientiertes PflegeAssessment Acute Care) nursing assessment tool, a standardised instrument completed by nursing staff at admission. The SPI ranges from 10 (complete dependence) to 40 (full independence) and shows strong convergent validity with the Extended Barthel Index (40–85% explained variance) [26] and predictive validity for 30-day mortality [27].

### Geriatric cohort definition

Patients were stratified into a geriatric and a non-geriatric cohort using a composite definition requiring both age ≥70 years and an adjusted HFRS ≥5 points (intermediate or high frailty risk). This dual criterion was chosen to operationalise a clinically meaningful geriatric phenotype that combines advanced age with demonstrable frailty, rather than relying on age alone. In the final sample, 2,053 patients (19.8%) met this definition.

### Statistical analysis

#### General approach

All analyses were conducted within a Bayesian inferential framework, which offers several advantages for this application: direct probability statements about parameters of interest, principled incorporation of prior evidence from both institutional historical data and published literature, and full posterior distributions enabling intuitive clinical interpretation [17,18]. All continuous covariates (age, HFRS, SPI) were standardised (z-scored) prior to model fitting.

#### Primary models

Both LOS and total costs were modelled using Bayesian generalised linear models with a Gamma distribution and log link function, reflecting the positive-valued, right-skewed nature of both outcomes [19–21]. Each model included dysphagia status (binary), geriatric cohort membership, their interaction, and all covariates listed above. The interaction term was included to test whether the proportional effect of dysphagia on outcomes differed between geriatric and non-geriatric patients. Model coefficients on the log scale represent log-ratios; exponentiated coefficients yield multiplicative effects.

#### Informative prior elicitation

Informative priors were constructed through a two-source synthesis. First, an independent institutional dataset comprising approximately 11,000 neurological inpatient cases from the same department (2013–2018) was analysed using frequentist Gamma-log generalised linear models with the same covariates. The resulting coefficient estimates and standard errors served as the primary source for prior means and variances. Second, published effect estimates from the literature were incorporated, including cost estimates from Attrill et al. [11] and Allen et al. [28], and intervention effect sizes from Carnaby et al. [14] and Dziewas et al. [15]. Prior distributions for each model parameter were specified as weighted averages of historical and literature evidence (approximately 60% historical, 30% literature, 10% clinical uncertainty buffer), with inflated standard errors to avoid overly informative priors.

#### MCMC sampling

Models were fitted using Hamiltonian Monte Carlo (HMC) via Stan, accessed through the brms package (version 2.22) in R (version 4.4.2) [22,23]. Each model used 4 parallel chains with 2,500 iterations per chain (1,000 warmup, 1,500 sampling), yielding 6,000 post-warmup draws in total. The target acceptance rate was set to 0.95 (adapt_delta). Chain convergence was assessed using the potential scale reduction factor (R^) and visual inspection of trace plots; all parameters achieved R^ < 1.01. Model fit was evaluated using posterior predictive checks (PPC).

#### Functional severity and intervention simulation models

To model the association between dysphagia severity and outcomes, two additional Gamma-log models were fitted within the dysphagic subgroup, regressing LOS and total costs on standardised FOIS scores (continuous), adjusted for all covariates. An ordinal FOIS model (cumulative logit, ordered categories 1–7) was additionally fitted to characterise the severity distribution.

Intervention effects were simulated using Bayesian posterior predictive simulation. For each dysphagic patient with available FOIS, we generated counterfactual predictions under hypothetical improvements of δFOIS = 1, 2, or 3 points on the raw FOIS scale (capped at 7). Results were summarised as mean differences with 95% credible intervals and as the posterior probability of any benefit (P[reduction > 0]). For the LOS-based model, day-based savings were additionally monetised using the institutional mean per-diem cost (€868.17, calculated as total departmental DRG revenue divided by total bed-days) to express LOS reductions in monetary terms; this conversion factor serves as a pragmatic approximation of the opportunity cost per bed-day and does not imply per-diem billing within the G-DRG system. All simulations were stratified by cohort.

#### Software

All analyses were performed in R (version 4.4.2) using brms (version 2.22) for Bayesian regression, posterior and tidybayes for posterior processing, and the tidyverse suite for data management. Stan served as the underlying probabilistic programming language for HMC sampling.

## Results

### Study population

A total of 15,390 inpatient cases were recorded in the Department of Neurology between January 2021 and December 2024. After excluding 467 cases with a length of stay of zero days (same-day discharges or data errors) and 789 mechanically ventilated patients, 14,134 cases remained. A further 3,759 cases were excluded due to missing covariate data (predominantly absent self-care index scores), yielding a final analytical sample of N = 10,375 cases (Figure 1).

Dysphagia was documented in 1,382 patients (13.3%), of whom 656 (47.5%) belonged to the non-geriatric and 726 (52.5%) to the geriatric cohort. The geriatric cohort (age ≥70 and adjusted HFRS ≥5) comprised 2,053 patients (19.8% of the sample). Among dysphagic patients, the mean Functional Oral Intake Scale (FOIS) score was 5.1 (SD 1.8), with geriatric patients showing lower functional oral intake (FOIS 4.8 vs. 5.5; Table 1).

**Table 1.** Descriptive statistics by cohort membership and dysphagia status. HFRS, Hospital Frailty Risk Score (adjusted for R13); SPI, self-care index; FOIS, Functional Oral Intake Scale (FOIS = 7 for non-dysphagic patients by definition). Costs in Euro (€).

| Cohort | n | Age<br>mean (SD) | HFRS<br>mean | SPI<br>mean | FOIS<br>mean | LOS<br>mean (SD) | LOS<br>med. | Cost<br>Mean (SD) | Cost<br>med. |
| --- | --- | --- | --- | --- | --- | --- | --- | --- | --- |
| Non-geriatric,<br>no dysphagia | 7,666 | 53.1 (18.2) | 2.7 | 38.3 | 7.0 | 6.6 (7.8) | 4 | €5,266<br>(4,746) | €3,499 |
| **Non-**<br>**geriatric,**<br>**Dysphagia** | 656 | 60.2 (12.1) | 8.6 | 31.3 | 5.5 | 14.7<br>(11.9) | 13 | €11,179<br>(12,303) | €8,857 |
| Geriatric,<br>no dysphagia | 1,327 | 80.0 (6.2) | 10.3 | 31.1 | 7.0 | 9.7 (8.2) | 7 | €7,063<br>(5,682) | €5,254 |
| **Geriatric,**<br>**Dysphagia** | 726 | 80.5 (6.2) | 15.4 | 23.5 | 4.8 | 18.8<br>(11.3) | 18 | €14,344<br>(9,600) | €11,715 |

Patients with dysphagia were older (mean age 70.8 vs. 56.9 years), had higher frailty burden (HFRS 12.3 vs. 3.9), and lower functional independence (SPI 27.1 vs. 37.0) than those without dysphagia. Unadjusted mean length of stay was approximately 2.5-fold longer in dysphagic patients across both cohorts (non-geriatric: 14.7 vs. 6.6 days; geriatric: 18.8 vs. 9.7 days). Unadjusted mean costs were similarly elevated (non-geriatric: €11,179 vs. €5,266; geriatric: €14,344 vs. €7,063).

### Model convergence and diagnostics

All five Bayesian models (primary LOS, primary cost, ordinal FOIS, FOIS-linked LOS, FOIS-linked cost) achieved satisfactory convergence. The maximum R^ across all parameters and models was 1.003, well below the conventional threshold of 1.01. Posterior predictive checks confirmed adequate model fit for both outcomes, with simulated data distributions closely matching the observed data (Supplementary Figures S1–S2).

### Dysphagia as independent predictor of LOS and costs

After adjustment for age, sex, Hospital Frailty Risk Score (R13-adjusted), stroke status, emergency admission, self-care index, and geriatric cohort membership, dysphagia remained a strong independent predictor of both outcomes (Table 2). Patients with documented dysphagia had a 46.5% longer length of stay (posterior ratio 1.465; 95% CrI 1.397–1.537) and 28.2% higher total case costs (posterior ratio 1.282; 95% CrI 1.213–1.354). Both credible intervals excluded 1.0, indicating strong posterior evidence for a non-zero effect.

**Table 2.** Adjusted posterior effect of dysphagia on length of stay and total case costs (Bayesian Gamma-log regression). Posterior ratios represent multiplicative effects on the outcome scale; CrI, credible interval.

| Outcome | Posterior Ratio | 95% CrI | Interpretation |
| --- | --- | --- | --- |
| Length of stay | 1.465 | 1.397–1.537 | +46.5% longer |
| Total case cost | 1.282 | 1.213–1.354 | +28.2% higher |

The discrepancy between the LOS and cost effects (46.5% vs. 28.2%) reflects the structure of the German G-DRG reimbursement system, in which total case costs are determined by diagnosis-related grouping, complication/comorbidity level, and length-of-stay trim points rather than by cumulative daily resource consumption. The two estimates address complementary economic perspectives: LOS quantifies the operational burden from the hospital’s perspective, where shorter stays enable higher case throughput and thus greater revenue under case-based reimbursement, whereas total case costs reflect the payer’s perspective, capturing the actual expenditure borne by health insurers.

### Geriatric stratification

The interaction between dysphagia and geriatric cohort membership was small but the 95% CrIs excluded 1.0 for both outcomes, and the effects were in opposite directions: dysphagia was associated with a proportionally smaller increase in LOS in geriatric patients (interaction ratio 0.908, 95% CrI 0.837–0.986) but a proportionally larger increase in total case costs (interaction ratio 1.096, 95% CrI 1.012–1.185). In practical terms, the LOS multiplier for dysphagia was approximately 1.33 in the geriatric cohort versus 1.47 in the non-geriatric cohort, while the cost multiplier was approximately 1.41 in the geriatric cohort versus 1.28 in the non-geriatric cohort.

Because geriatric patients have substantially higher baseline values for both LOS and costs, even the slightly attenuated LOS ratio still translates into a larger absolute burden. Applied to a geriatric baseline of 9.7 days, the geriatric-specific LOS ratio yields approximately 3.2 additional days, compared to 3.1 additional days in the non-geriatric cohort (baseline 6.6 days). For costs, the combination of a higher proportional effect and a higher baseline amplifies the absolute difference: the cost increment is approximately €2,900 in the geriatric cohort versus €1,500 in the non-geriatric cohort.

### Functional severity and dose–response relationship

The ordinal FOIS model confirmed a dose–response relationship between functional swallowing severity and both outcomes. Each unit decrease in FOIS (indicating greater dysphagia severity) was associated with progressively longer hospital stays and higher costs. The FOIS-linked models estimated the posterior expected mean FOIS at 6.77 (95% CrI 4.91–6.99); this represents the population-level model expectation across all patients (including non-dysphagic patients assigned FOIS = 7), whereas the observed mean among dysphagic patients only was 5.14 (SD 1.77). The wide credible interval encompasses both values, confirming that the model adequately captures the heterogeneity in swallowing function across the full severity spectrum.

### Intervention simulation

Bayesian posterior predictive simulation quantified the expected reductions in LOS and costs associated with hypothetical improvements in swallowing function (δFOIS = 1, 2, or 3 scale points). Table 3 summarises the results stratified by cohort.

**Table 3.**
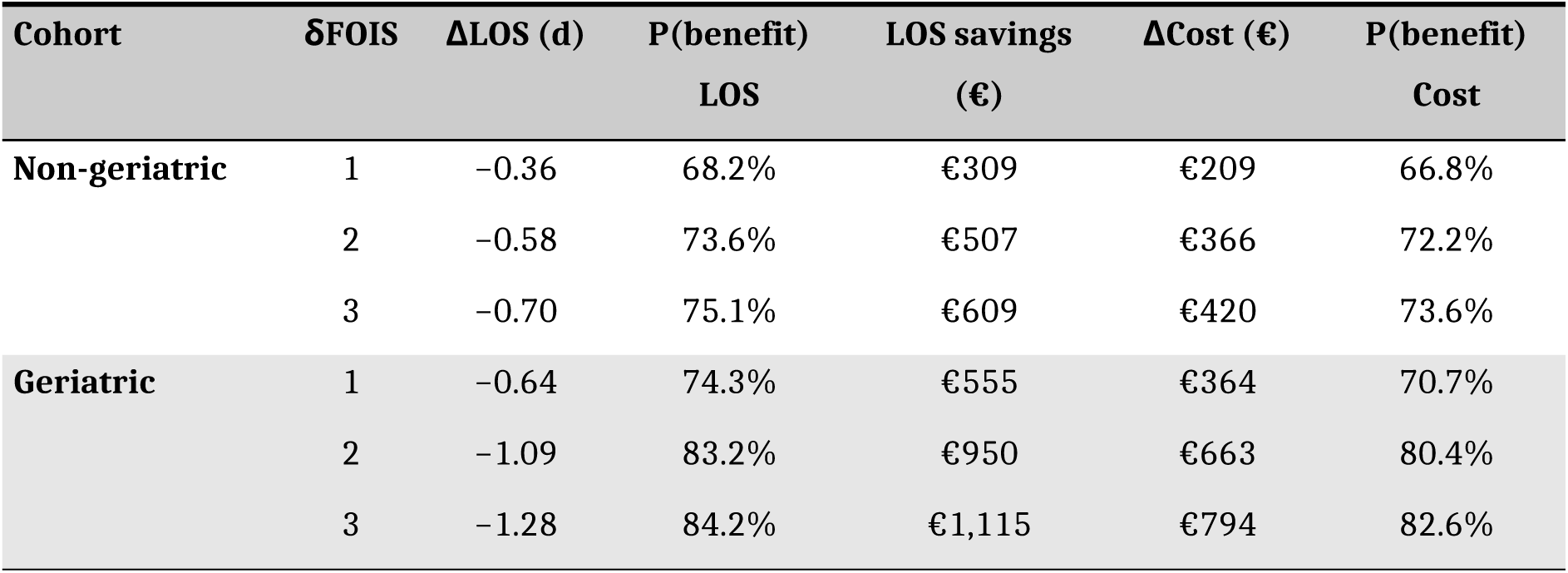
Posterior predictive simulation of FOIS improvements on LOS and costs by cohort. δFOIS, simulated improvement in Functional Oral Intake Scale score; ΔLOS, mean change in days; P(benefit), posterior probability of a favourable outcome; LOS savings, monetised LOS reduction (institutional per-diem rate €868.17); ΔCost, mean change from the direct cost model (G-DRG).

In the geriatric cohort, a one-point FOIS improvement was associated with a mean LOS reduction of 0.64 days (per-diem equivalent: €555) with a 74.3% posterior probability of benefit. At δFOIS = 3, the mean LOS reduction increased to 1.28 days (€1,115 per case) with an 84.2% probability of benefit. Geriatric patients consistently showed higher absolute savings and higher probabilities of benefit compared to non-geriatric patients, reflecting their higher baseline resource utilisation.

The direct cost model yielded similar but slightly attenuated probabilities of benefit (82.6% at δFOIS = 3 in the geriatric cohort). As outlined above, the two models address complementary economic perspectives: LOS-based savings reflect the hospital’s throughput potential, whereas direct cost savings capture the payer’s burden within the G-DRG system. Both approaches converge on the conclusion that functional improvements in swallowing carry a high probability of generating cost savings, particularly in geriatric patients.

## Discussion

To our knowledge, this study provides the first comprehensive, Bayesian-adjusted analysis of the economic burden of neurogenic dysphagia in a large German neurological inpatient population. Three principal findings emerged. First, dysphagia was independently associated with a 46.5% increase in length of stay and a 28.2% increase in total case costs after adjustment for age, sex, frailty, functional status, stroke diagnosis, and admission type, indicating that it is a substantial and independent cost driver and not simply a marker of disease severity or functional deficit. As such, dysphagia emerges as a viable target for interventions aiming at reducing resource consumption. Second, small but credible interactions between dysphagia and geriatric status revealed a slightly attenuated LOS effect but a slightly amplified cost effect in geriatric patients, with the absolute economic burden remaining unambiguously larger in the geriatric cohort due to higher baseline resource utilisation. Third, Bayesian posterior predictive simulation demonstrated that improvements in functional swallowing status carry a high probability of generating cost savings, particularly in geriatric patients.

### Dysphagia as an independent cost driver

The adjusted posterior ratios in our study (LOS: 1.465, 95% CrI 1.397–1.537; cost: 1.282, CrI 1.213–1.354) are notably lower than most previously reported estimates. The meta-analysis by Attrill et al. [12] reported an approximately 40% increase in costs, but this estimate was derived primarily from unadjusted studies. Patel et al. [13], using inverse probability weighting in a US sample of 88 million records, found a 76% increase in length of stay, but did not adjust for frailty or functional status. Our lower cost estimate likely reflects the absorption of confounding variance by the HFRS and self-care index, both of which are strongly correlated with dysphagia and independently drive costs. The fact that a substantial and credibly non-zero effect persists after this comprehensive adjustment strengthens the interpretation that dysphagia acts as an independent cost driver rather than merely being a proxy for overall disease severity.

Critically, the choice of adjustment variables is not merely assumption-driven. In a serial mediation analysis conducted on an independent institutional cohort (2014–2018, n = 1,116; Werner CJ et al., submitted), the pathway from chronological age to swallowing impairment was fully mediated through frailty (HFRS) and functional status (SPI), with no significant direct effect of age on dysphagia severity (β = 0.016, p = 0.607). This confirms that the present model adjusts for the empirically validated upstream determinants of dysphagia, not merely for plausible confounders. The residual cost effect therefore cannot easily be attributed to unmeasured confounding along the age–frailty–dysphagia pathway. Instead, it suggests that once dysphagia is established - regardless of its origins in the frailty–disability cascade - it generates additional resource consumption through its own specific mechanisms, including e.g., aspiration management, dietary modification, prolonged monitoring, and the treatment of complications such as hospital-acquired pneumonia. However, as both analyses are observational, causal inferences should be drawn with appropriate caution.

### Comparison with German cost data

The most directly comparable German study is that of Labeit et al. [13], who reported €2,554 in additional costs for severe post-stroke dysphagia (FEDSS = 6) in 674 patients. Several differences may explain divergent findings: Labeit et al. used ordinary least squares regression rather than a Gamma-log model [19], restricted their analysis to stroke patients, did not adjust for frailty, and did not exclude mechanically ventilated patients, whose extreme costs may inflate estimates at the severe end of the spectrum, even after inclusion of ventilation time as a covariate.

Our approach complements theirs through a broader population (N = 10,375, all neurological diagnoses), exclusion of ventilated patients, comprehensive covariate adjustment including HFRS and SPI, and Bayesian inference.

### Differential geriatric interaction across outcomes

The opposing interaction directions - a slightly attenuated LOS effect but amplified cost effect in geriatric patients - deserve interpretation. This divergence is plausible within the G-DRG framework. Geriatric patients with dysphagia may accumulate higher case costs through complexity-driven grouping (higher CC levels, geriatric complex treatment codes) even when LOS is not proportionally increased to the same degree as in non-geriatric patients. Conversely, the slightly lower LOS interaction may reflect systematic care differences: geriatric patients with dysphagia may be transferred earlier to rehabilitation or geriatric follow-up facilities, capping acute-care LOS while total case costs continue to accrue through the DRG grouping mechanism. Importantly, the absolute economic burden of dysphagia remains unambiguously larger in geriatric patients due to their higher baseline resource utilisation: the higher cost ratio applied to a higher baseline (€7,063 vs. €5,266 in non-dysphagic patients) yields an absolute cost increment of approximately €2,900 per case in the geriatric cohort versus €1,500 in the non-geriatric cohort.

These findings have direct implications for resource allocation. Because dysphagia drives meaningful cost increases in both cohorts, albeit through slightly different mechanisms, dysphagia management is relevant across all age groups and frailty levels, not only in the geriatric population where it has traditionally received the most attention [8]. The higher absolute savings potential in geriatric patients provides a strong economic argument for prioritising dysphagia screening and intervention in this subgroup, however, consistent with the European Society for Swallowing Disorders’ recommendation to treat oropharyngeal dysphagia as a geriatric syndrome warranting systematic screening [8].

### From functional improvement to cost savings

The intervention simulation represents, to our knowledge, a novel contribution to the dysphagia literature. A systematic review by Marin et al. [16] identified few formally cost-effective dysphagia intervention across the published literature, highlighting an important evidence gap between clinical efficacy and economic justification. Our approach bridges this gap by linking improvements on the FOIS scale, an outcome that is routinely assessed and clinically interpretable [24], directly to probabilistic estimates of cost savings via Bayesian posterior predictive simulation.

The finding that a one-point FOIS improvement in geriatric patients yields a 74.3% posterior probability of cost savings (mean €555 per case based on LOS reduction) may be clinically actionable. For context, a one-point FOIS improvement corresponds, for example, to a transition from complete tube dependent feeding (nothing by mouth) to tube dependent feeding with minimal attempts of food and liquid, or from pureed diet alone to pureed and soft food diet. Such improvements are achievable through established interventions including behavioural swallowing therapy [14], texture modification [29], and neurostimulatory approaches such as pharyngeal electrical stimulation [15]. At three FOIS points of improvement, the probability of benefit exceeds 84% with mean savings of €1,115 per case. These estimates are deliberately therapy-agnostic: they provide a ceiling value against which specific interventions can be evaluated in future cost-effectiveness analyses.

That the probability of benefit is consistently high under both the hospital perspective (LOS-based: 84.2%) and the payer perspective (direct cost: 82.6% at δFOIS = 3 in the geriatric cohort) strengthens the economic argument, as it suggests potential gains for hospitals and payers alike.

### Strengths and limitations

Several strengths of this study deserve emphasis. The large sample size (N = 10,375) encompasses the full spectrum of neurological diagnoses seen in a university hospital setting, enhancing generalisability beyond the stroke-only populations studied previously [12,13]. The Bayesian framework with informative priors from an independent historical dataset (2013–2018, approximately 11,000 cases) and published literature enables direct probabilistic inference, avoids the pitfalls of null-hypothesis significance testing, and provides decision-relevant output for health economic policy [18]. The adjustment of the Hospital Frailty Risk Score for the R13 (dysphagia) ICD-10 code [25] eliminates a potential source of collinearity that has not been addressed in prior studies. The exclusion of mechanically ventilated patients provides a cleaner estimate of the dysphagia-specific cost effect, uncontaminated by the extreme costs associated with intensive care.

Several limitations should be acknowledged. First, this is a single-centre study, though the G-DRG system provides a standardised reimbursement framework across German hospitals [30], limiting the scope for idiosyncratic cost structures. Second, the composite dysphagia classification (FEES documentation and/or ICD-10 R13 coding) may differ in sensitivity and specificity from instrumental assessment alone. Third, the exclusion of 3,759 cases with missing covariates (predominantly absent SPI scores) may introduce selection bias, though missingness likely reflects short-stay patients or administrative factors rather than systematic clinical differences. Fourth, the intervention simulation is based on observational dose–response modelling; the simulated FOIS improvements represent expected values conditional on improvement, not estimates of any particular intervention’s efficacy.

### Clinical implications and future directions

Our findings have three practical implications for clinical care and health policy. First, they support the implementation of systematic dysphagia screening in neurological inpatients, not only for clinical reasons but also for economic ones. Given the large proportional cost effect (28.2%) and its independence from frailty and functional status, early identification and management of dysphagia represents a potentially modifiable target for hospital cost reduction. Second, the finding that dysphagia drives substantial cost increases in both geriatric and non-geriatric cohorts, despite small differences in the proportional effect, argues against restricting dysphagia-focused resource allocation to geriatric patients alone. Third, the probabilistic savings estimates from our intervention simulation provide a framework for future cost-effectiveness analyses: our models suggest that any intervention achieving a one-point FOIS improvement would carry a greater than 70% posterior probability of being cost-saving, though this estimate requires prospective validation.

Future studies should validate these findings in multicentre datasets and in other healthcare systems, and should prospectively link specific dysphagia interventions to FOIS-based cost savings using the Bayesian simulation framework developed here. The extension to long-term costs (post-discharge rehabilitation, readmission) and quality-of-life outcomes would further strengthen the economic case for dysphagia management in neurological populations.

## Conclusion

Neurogenic dysphagia is an independent and substantial driver of hospital length of stay and costs in neurological inpatients, with a 46.5% increase in length of stay and 28.2% increase in total case costs persisting after comprehensive Bayesian adjustment for frailty, functional status, and other confounders. Small but credible differences in the proportional effect between geriatric and non-geriatric patients (slightly attenuated for LOS, slightly amplified for costs) are consistent with the mechanics of the G-DRG system and do not diminish the clinical relevance of dysphagia in either cohort. The absolute economic burden remains unambiguously larger in geriatric patients. Bayesian posterior predictive simulation indicates that improvements in swallowing function carry a high probability of generating cost savings, with geriatric patients standing to benefit most. These findings support the characterisation of dysphagia as a modifiable economic target and provide decision-relevant estimates for health policy and intervention planning.

## Acknowledgments

Not applicable.

## Data Availability

The datasets analysed in the current study are not publicly available as they contain confidential institutional reimbursement data subject to data protection regulations.

## Supplementary Materials

The following supplementary materials are available online:

**Supplementary Figure S1.** Posterior predictive checks for the primary LOS model.

**Supplementary Figure S2.** Posterior predictive checks for the primary cost model.

**Supplementary Figure S3.** Posterior density distributions of effect ratios for all covariates in the primary length of stay and cost models.

**Supplementary Table S1.** Full posterior coefficient estimates, prior specifications, and convergence diagnostics for all Bayesian models.

## References

[1] Martino R, Foley N, Bhogal S, Diamant N, Speechley M, Teasell R. Dysphagia After Stroke: Incidence, Diagnosis, and Pulmonary Complications. Stroke 2005;36:2756–63. 10.1161/01.STR.0000190056.76543.eb.

[2] Kalf JG, De Swart BJM, Bloem BR, Munneke M. Prevalence of oropharyngeal dysphagia in Parkinson’s disease: A meta-analysis. Parkinsonism & Related Disorders 2012;18:311–5. 10.1016/j.parkreldis.2011.11.006.

[3] Takizawa C, Gemmell E, Kenworthy J, Speyer R. A Systematic Review of the Prevalence of Oropharyngeal Dysphagia in Stroke, Parkinson’s Disease, Alzheimer’s Disease, Head Injury, and Pneumonia. Dysphagia 2016;31:434–41. 10.1007/s00455-016-9695-9.

[4] Putri AR, Chu Y-H, Chen R, Chiang K-J, Banda KJ, Liu D, et al. Prevalence of swallowing disorder in different dementia subtypes among older adults: a meta-analysis. Age and Ageing 2024;53:afae037. 10.1093/ageing/afae037.

[5] Banda KJ, Chu H, Kang XL, Liu D, Pien L-C, Jen H-J, et al. Prevalence of dysphagia and risk of pneumonia and mortality in acute stroke patients: a meta-analysis. BMC Geriatr 2022;22:420. 10.1186/s12877-022-02960-5.

[6] Carrión S, Cabré M, Monteis R, Roca M, Palomera E, Serra-Prat M, et al. Oropharyngeal dysphagia is a prevalent risk factor for malnutrition in a cohort of older patients admitted with an acute disease to a general hospital. Clinical Nutrition 2015;34:436–42. 10.1016/j.clnu.2014.04.014.

[7] Rivelsrud MC, Hartelius L, Bergström L, Løvstad M, Speyer R. Prevalence of Oropharyngeal Dysphagia in Adults in Different Healthcare Settings: A Systematic Review and Meta-analyses. Dysphagia 2023;38:76–121. 10.1007/s00455-022-10465-x.

[8] Baijens LW, Clavé P, Cras P, Ekberg O, Forster A, Kolb G, et al. European Society for Swallowing Disorders – European Union Geriatric Medicine Society white paper: oropharyngeal dysphagia as a geriatric syndrome. CIA 2016;Volume 11:1403–28. 10.2147/CIA.S107750.

[9] Melgaard D, Rodrigo-Domingo M, Mørch M. The Prevalence of Oropharyngeal Dysphagia in Acute Geriatric Patients. Geriatrics 2018;3:15. 10.3390/geriatrics3020015.

[10] Warnecke T, Dziewas R, Wirth R, Bauer JM, Prell T. Dysphagia from a neurogeriatric point of view: Pathogenesis, diagnosis and management. Z Gerontol Geriatr 2019;52:330–5. 10.1007/s00391-019-01563-x.

[11] Attrill S, White S, Murray J, Hammond S, Doeltgen S. Impact of oropharyngeal dysphagia on healthcare cost and length of stay in hospital: a systematic review. BMC Health Serv Res 2018;18:594. 10.1186/s12913-018-3376-3.

[12] Patel DA, Krishnaswami S, Steger E, Conover E, Vaezi MF, Ciucci MR, et al. Economic and survival burden of dysphagia among inpatients in the United States. Dis Esophagus 2018;31:dox131. 10.1093/dote/dox131.

[13] Labeit B, Kremer A, Muhle P, Claus I, Warnecke T, Dziewas R, et al. Costs of post-stroke dysphagia during acute hospitalization from a health-insurance perspective. European Stroke Journal 2023;8:361–9. 10.1177/23969873221147740.

[14] Carnaby G, Hankey GJ, Pizzi J. Behavioural intervention for dysphagia in acute stroke: a randomised controlled trial. The Lancet Neurology 2006;5:31–7. 10.1016/S1474-4422(05)70252-0.

[15] Dziewas R, Stellato R, Van Der Tweel I, Walther E, Werner CJ, Braun T, et al. Pharyngeal electrical stimulation for early decannulation in tracheotomised patients with neurogenic dysphagia after stroke (PHAST-TRAC): a prospective, single-blinded, randomised trial. The Lancet Neurology 2018;17:849–59. 10.1016/S1474-4422(18)30255-2.

[16] Marin S, Ortega O, Serra-Prat M, Valls E, Pérez-Cordón L, Clavé P. Economic Evaluation of Clinical, Nutritional and Rehabilitation Interventions on Oropharyngeal Dysphagia after Stroke: A Systematic Review. Nutrients 2023;15:1714. 10.3390/nu15071714.

[17] Briggs AH, Claxton K, Sculpher MJ. Decision modelling for health economic evaluation. Oxford: Oxford University Press; 2006.

[18] Baio G. Bayesian Methods in Health Economics. 0 ed. Chapman and Hall/CRC; 2012. 10.1201/b13099.

[19] Manning WG, Mullahy J. Estimating log models: to transform or not to transform? Journal of Health Economics 2001;20:461–94. 10.1016/S0167-6296(01)00086-8.

[20] Malehi AS, Pourmotahari F, Angali KA. Statistical models for the analysis of skewed healthcare cost data: a simulation study. Health Econ Rev 2015;5:11. 10.1186/s13561-015-0045-7.

[21] Nixon RM, Thompson SG. Parametric modelling of cost data in medical studies. Statistics in Medicine 2004;23:1311–31. 10.1002/sim.1744.

[22] Bürkner P-C. brms: An R Package for Bayesian Multilevel Models Using Stan. J Stat Soft 2017;80. 10.18637/jss.v080.i01.

[23] Bürkner P-C. Advanced Bayesian Multilevel Modeling with the R Package brms. The R Journal 2018;10:395. 10.32614/RJ-2018-017.

[24] Crary MA, Mann GDC, Groher ME. Initial Psychometric Assessment of a Functional Oral Intake Scale for Dysphagia in Stroke Patients. Archives of Physical Medicine and Rehabilitation 2005;86:1516–20. 10.1016/j.apmr.2004.11.049.

[25] Gilbert T, Neuburger J, Kraindler J, Keeble E, Smith P, Ariti C, et al. Development and validation of a Hospital Frailty Risk Score focusing on older people in acute care settings using electronic hospital records: an observational study. The Lancet 2018;391:1775–82. 10.1016/S0140-6736(18)30668-8.

[26] Suter-Riederer S, Schwarz J, Imhof L, Petry H. Vergleichbarkeit von ergebnisorientiertem Pflegeassessment (ePA_AC) und Erweitertem Barthel Index (EBI). Neurologie & Rehabilitation 2014;20:24–30.

[27] Koch D, Schuetz P, Haubitz S, Kutz A, Mueller B, Weber H, et al. Improving the post-acute care discharge score (PACD) by adding patients’ self-care abilities: A prospective cohort study. PLoS One 2019;14:e0214194. 10.1371/journal.pone.0214194.

[28] Allen J, Greene M, Sabido I, Stretton M, Miles A. Economic costs of dysphagia among hospitalized patients. The Laryngoscope 2020;130:974–9. 10.1002/lary.28194.

[29] Dziewas R, Allescher H-D, Aroyo I, Bartolome G, Beilenhoff U, Bohlender J, et al. Diagnosis and treatment of neurogenic dysphagia – S1 guideline of the German Society of Neurology. Neurol Res Pract 2021;3:23. 10.1186/s42466-021-00122-3.

[30] Busse R, Geissler A, Aaviksoo A, Cots F, Hakkinen U, Kobel C, et al. Diagnosis related groups in Europe: moving towards transparency, efficiency, and quality in hospitals? BMJ 2013;346:f3197–f3197. 10.1136/bmj.f3197.

