## Supplemental Materials for "Neurogenic dysphagia as an independent driver of hospital length of stay and costs: *a Bayesian analysis with geriatric stratification and intervention simulation*"

Werner CJ et al.

**Supplementary Figure S1**

*Posterior Predictive Check for Length of Stay Model*

Posterior predictive check (PPC) comparing observed length of stay data (black line) with replicated datasets from the posterior predictive distribution (light blue lines). The alignment between observed and replicated data indicates adequate model fit.

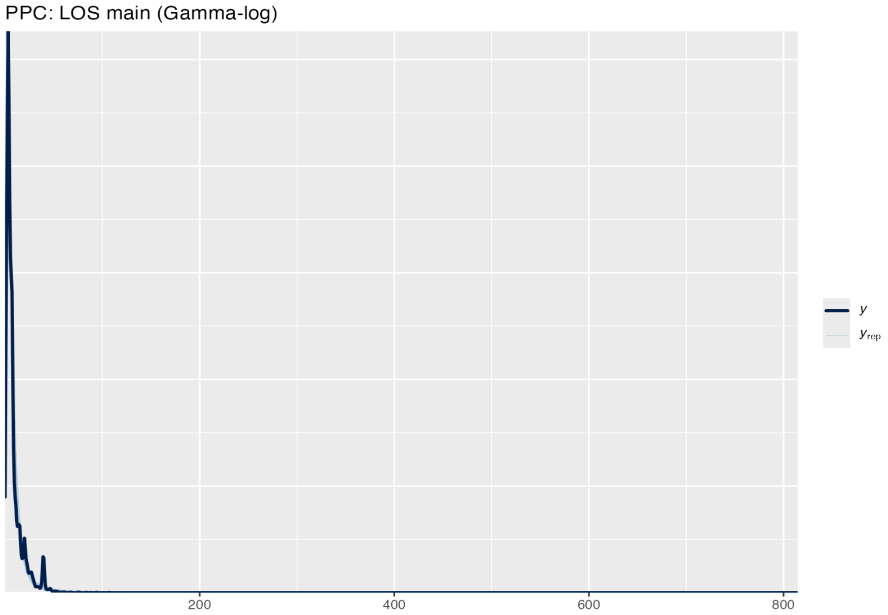


**Supplementary Figure S2**

*Posterior Predictive Check for Cost Model*

Posterior predictive check (PPC) comparing observed health service costs (black line) with replicated cost datasets from the posterior predictive distribution (light blue lines). The comparison demonstrates appropriate model calibration to the observed cost distribution.

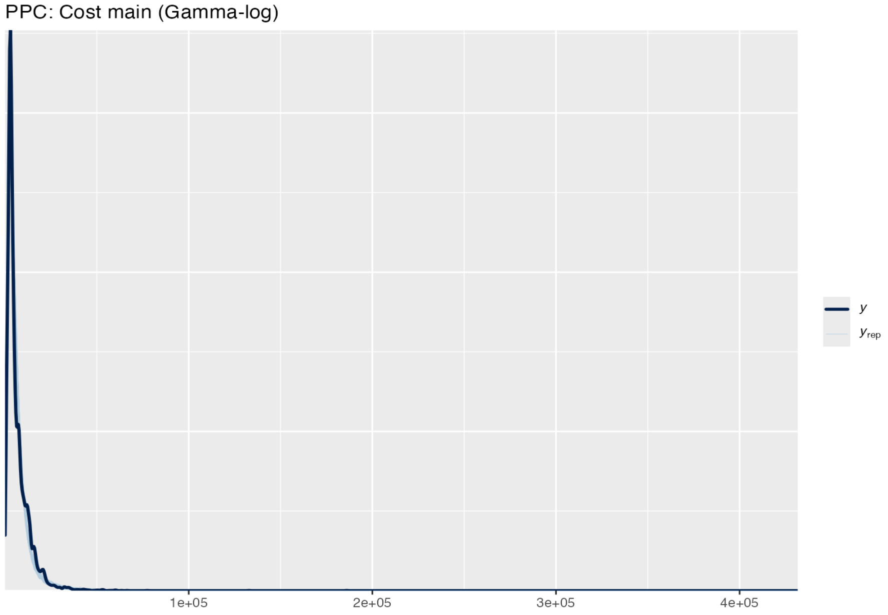


**Supplementary Figure S3**

*Posterior Density Estimates of Effect Ratios*

Posterior density distributions of exponentiated regression coefficients (effect ratios) for both the length of stay and cost models. Density curves are shown for each covariate, with posterior means indicated by vertical dashed lines.

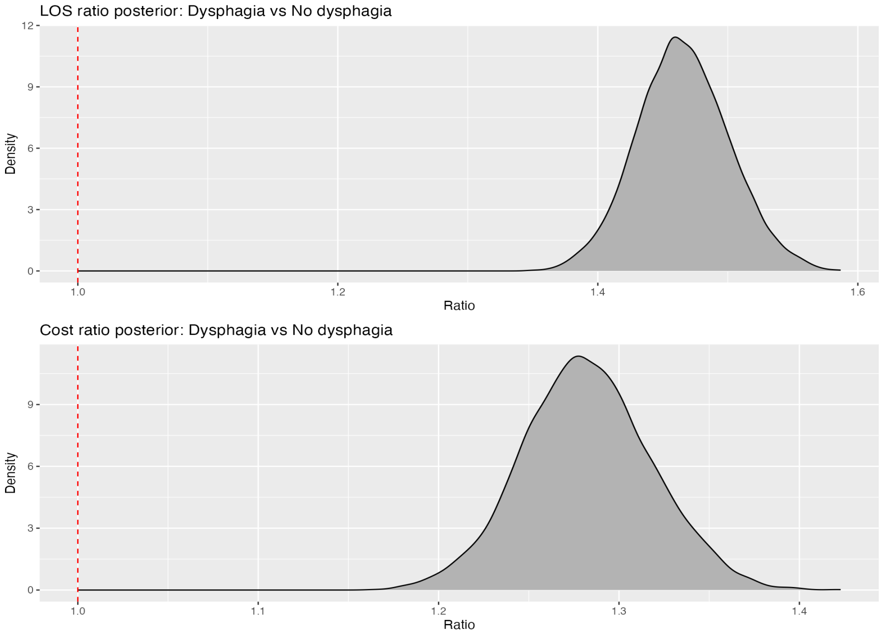


**Supplementary Table S1**

**Panel A: Prior Specifications**

| Model | Parameter | Prior distribution | Source |
|---|---|---|---|
| LOS primary | Intercept | Normal(2.079, 0.300) | Historical GLM |
| | Dysphagia (Yes) | Normal(0.520, 0.180) | Synthesised |

| | | | |
|---|---|---|---|
| | Age (std) | Normal(μ_h, 1.5×SE_h) | Historical |
| | Sex (female) | Normal(μ_h, 1.5×SE_h) | Historical |
| | Stroke | Normal(μ_h, 1.5×SE_h) | Historical |
| | Emergency | Normal(μ_h, 1.5×SE_h) | Historical |
| | HFRS (std) | Normal(μ_h, 1.5×SE_h) | Historical |
| | SPI (std) | Normal(μ_h, 1.5×SE_h) | Historical |
| | Geriatric cohort | Normal(0, 0.5) | Weakly informative |
| | Dysphagia × Geriatric | Normal(0, 0.5) | Weakly informative |
| | Shape (α) | Exponential(1) | Default |
| Cost primary | Intercept | Normal(8.250, 0.300) | Historical GLM |
| | Dysphagia (Yes) | Normal(0.480, 0.180) | Synthesised |
| | Covariates | As LOS primary model | Historical |
| | Shape (α) | Exponential(1) | Default |
| FOIS ordinal | Thresholds, covariates | Flat (brms defaults) | Non-informative |
| LOS ~ FOIS | Intercept | Normal(2.079, 0.300) | Historical GLM |
| | FOIS (std) | Normal(−0.200, 0.120) | Literature synth. |
| | Covariates | As LOS primary model | Historical |
| | Shape (α) | Exponential(1) | Default |
| Cost ~ FOIS | Intercept | Normal(8.250, 0.300) | Historical GLM |
| | FOIS (std) | Normal(−0.200, 0.120) | Literature synth. |
| | Covariates | As Cost primary model | Historical |
| | Shape (α) | Exponential(1) | Default |

**Panel B: Full Posterior Estimates**

**B1. LOS Primary Model**

| Parameter | log(β) | Est.Error | Ratio (exp) | 95% CrI [lower–upper] |
|---|---|---|---|---|
| Intercept | 2.0758 | 0.0131 | 7.971 | 7.773–8.178 |
| Dysphagia (Yes) | 0.3820 | 0.0241 | 1.465 | 1.397–1.537 |
| Geriatric | −0.1642 | 0.0263 | 0.849 | 0.807–0.893 |
| Age (std) | −0.0099 | 0.0075 | 0.990 | 0.976–1.005 |
| Sex (female) | −0.1201 | 0.0122 | 0.887 | 0.866–0.909 |
| Stroke | −0.0126 | 0.0150 | 0.987 | 0.959–1.017 |
| Emergency | −0.0595 | 0.0137 | 0.942 | 0.917–0.968 |
| HFRS (std) | 0.4692 | 0.0087 | 1.599 | 1.572–1.626 |
| SPI (std) | −0.0242 | 0.0084 | 0.976 | 0.960–0.992 |
| Dysphagia × Geriatric | −0.0963 | 0.0417 | 0.908 | 0.837–0.986 |

**B2. Cost Primary Model**

| Parameter | log(β) | Est.Error | Ratio (exp) | 95% CrI [lower–upper] |
|---|---|---|---|---|

| | | | | |
|---|---|---|---|---|
| Intercept | 8.7444 | 0.0123 | 6275.19 | 6127.05–6428.51 |
| Dysphagia (Yes) | 0.2482 | 0.0279 | 1.282 | 1.213–1.354 |
| Geriatric | −0.1621 | 0.0237 | 0.850 | 0.812–0.891 |
| Age (std) | −0.0590 | 0.0068 | 0.943 | 0.930–0.956 |
| Sex (female) | −0.0893 | 0.0111 | 0.915 | 0.895–0.934 |
| Stroke | 0.2564 | 0.0142 | 1.292 | 1.257–1.330 |
| Emergency | −0.2792 | 0.0126 | 0.756 | 0.738–0.775 |
| HFRS (std) | 0.4030 | 0.0080 | 1.496 | 1.473–1.521 |
| SPI (std) | −0.0204 | 0.0074 | 0.980 | 0.966–0.994 |
| Dysphagia × Geriatric | 0.0916 | 0.0399 | 1.096 | 1.012–1.185 |

**B3. LOS ~ FOIS Model (dysphagic subgroup)**

| Parameter | log(β) | Est.Error | Ratio (exp) | 95% CrI [lower–upper] |
|---|---|---|---|---|
| Intercept | 1.9902 | 0.0372 | 7.317 | 6.801–7.875 |
| FOIS (std) | −0.0446 | 0.0099 | 0.956 | 0.938–0.975 |
| Age (std) | 0.0354 | 0.0285 | 1.036 | 0.981–1.095 |
| Sex (female) | −0.1092 | 0.0376 | 0.897 | 0.832–0.963 |
| Stroke | −0.0500 | 0.0456 | 0.951 | 0.869–1.039 |
| Emergency | −0.0438 | 0.0454 | 0.957 | 0.874–1.044 |
| HFRS (std) | 0.2302 | 0.0175 | 1.259 | 1.217–1.303 |
| SPI (std) | −0.0644 | 0.0198 | 0.938 | 0.902–0.975 |

**B4. Cost ~ FOIS Model (dysphagic subgroup)**

| Parameter | log(β) | Est.Error | Ratio (exp) | 95% CrI [lower–upper] |
|---|---|---|---|---|
| Intercept | 8.2372 | 0.0389 | 3779.13 | 3504.63–4078.54 |
| FOIS (std) | −0.0407 | 0.0101 | 0.960 | 0.942–0.979 |
| Age (std) | 0.0145 | 0.0295 | 1.015 | 0.959–1.073 |
| Sex (female) | −0.1055 | 0.0380 | 0.900 | 0.835–0.970 |
| Stroke | 0.4178 | 0.0442 | 1.519 | 1.393–1.655 |
| Emergency | −0.0547 | 0.0450 | 0.947 | 0.867–1.034 |
| HFRS (std) | 0.2307 | 0.0179 | 1.260 | 1.217–1.305 |
| SPI (std) | −0.0370 | 0.0198 | 0.964 | 0.927–1.001 |

**Panel C: Convergence Diagnostics**

| Model | Max $\hat{R}$ |
|---|---|
| LOS (primary) | 1.001 |
| Cost (primary) | 1.003 |
| FOIS (ordinal) | 1.002 |
| LOS ~ FOIS | 1.002 |

| | |
|---|---|
| Cost ~ FOIS | 1.002 |

**Note:**

MCMC convergence was assessed using the $\hat{R}$ statistic, with values below 1.01 indicating effective convergence. All models were fitted using 4 independent Markov chains with 2,500 iterations each, including 1,000 warmup iterations per chain, with adapt_delta set to 0.95 for improved sampling efficiency. μ_h and SE_h denote the mean and standard error of covariates from the historical dataset. Standardised (std) covariates were centred on the historical mean and scaled by the historical standard deviation.
